# Improving effectiveness of different deep learning-based models for detecting COVID-19 from computed tomography (CT) images

**DOI:** 10.1101/2020.06.12.20129643

**Authors:** Erdi Acar, Engin Şahin, İhsan Yılmaz

## Abstract

Computerized Tomography (CT) has a prognostic role in the early diagnosis of COVID-19 due to it gives both fast and accurate results. This is very important to help decision making of clinicians for quick isolation and appropriate patient treatment. In this study, we combine methods such as segmentation, data augmentation and the generative adversarial network (GAN) to improve the effectiveness of learning models. We obtain the best performance with 99% accuracy for lung segmentation. Using the above improvements we get the highest rates in terms of accuracy (99.8%), precision (99.8%), recall (99.8%), f1-score (99.8%) and roc acu (99.9979%) with deep learning methods in this paper. Also we compare popular deep learning-based frameworks such as VGG16, VGG19, Xception, ResNet50, ResNet50V2, DenseNet121, DenseNet169, InceptionV3 and InceptionResNetV2 for automatic COVID-19 classification. The DenseNet169 amongst deep convolutional neural networks achieves the best performance with 99.8% accuracy. The second-best learner is InceptionResNetV2 with accuracy of 99.65%. The third-best learner is Xception and InceptionV3 with accuracy of 99.60%.

## 1 Introduction

Coronavirus disease (COVID-19) has led to huge public health problem in the international community since it is rapidly spreading all over the World. Although polymerase chain reaction (PCR) test is standard for confirming COVID-19 positive patients, medical imaging such as X-ray and non-contrast computed tomography (CT) plays an important role in COVID-19 detection. Since COVID-19 can be determined by the presence of lung ground-glass opacities on CT images, which is clearer and more precise according to X-ray images, in early stages, diagnosis of COVID-19 from CT will help decision making of clinicians for quick isolation and appropriate patient treatment [1]. However chest CT images take more time for the specialists to diagnose. Therefore, it is important to use CT images for automated diagnosis of COVID-19. For this aim it is purposed deep learning-based frameworks [2].

Chen et al. [3] determine COVID-19 or non-COVID-19 from CT images including 51 COVID-19 patients and 55 patients with other diseases using UNet++ segmentation model. The results of COVID-19 classification are 95.2% (accuracy), 100% (sensitivity), and 93.6% (specificity). A U-Net and 3D CNN models are used for lung segmentation and diagnosing of COVID-19, respectively, in Ref. [4]. The results of the model are 90.7% (sensitivity), 91.1% (specificity), and 0.959 (AUC). Jin et al. [5] determine COVID-19 or non-COVID-19 from CT images including 496 COVID-19 positive cases and 1385 negative cases using 2D CNN based model. The results of the model are 94.1% (sensitivity), 95.5% (specificity), and 0.979 (AUC). Also Jin et al. [6] propose ResNet50 for classification and UNet++ for segmentation using CT images of 1136 cases (i.e., 723 COVID-19 positives, and 413 COVID-19 negatives). The results of the model are 97.4% (sensitivity) and 92.2% (specificity). Most of the previous works on COVID-19 are given in Ref. [7].

The main contributions of this paper can be summarized as:

− It is difficult to identify COVID-19 in CT images due to infections caused by COVID-19 often occur in small regions of the lungs [1] and the large variation in both position and shape across different patients [8]. Therefore we will use segmentation model based on bidirectional ConvLSTM U-Net [9] and graph-cut image processing [10] to improve the effectiveness of learning models.
− To improve the effectiveness of learning models we use data augmentation operations such as Random distortion, Flip, Rotate and Zoom.
− Deep learning algorithms require a large data set for training. If the model is trained with a small data set, the model becomes overfitting [8]. For this aim we will use the generative adversarial network (GAN) to generate more images and overcome the overfitting problem.
− Due to the above improvements, we achieved a high success of 99.80% in detecting COVID-19.

The rest of the paper is organized as follows. In Section 2, the materials and methods used in the study are presented. Section 3 presents the results of different analyzes for different deep learning algorithms in the proposed frameworks. The comparisons with the other related studies and the discussion of the results are given in Sec. 4.

## 2 Material and methods

### 2.1 Dataset

We collect the CT images from different open sources in Refs. [11, 12]. Some of the CT images collected from the data sources are ignored due to repetition or high correlation problems in the CT images on the source. The chest CT images in our dataset consist of 1232 COVID-19 and 1668 healthy images. From the CT images in our dataset, 986 COVID-19 images and 1334 healthy images are randomly selected for training set. 99 COVID-19 images and 133 healthy images are randomly selected for validation set. 246 COVID-19 images and 334 healthy images are randomly selected for testing set.

### 2.2 Improving the data

This section describes the steps (lung segmentation, pre-processing and generative adversarial network) which are carried out to make training better.

#### 2.2.1 Lung segmentation

BConvLSTM U-Net [9] architecture is used for segmentation. 1606 training size and 1606 mask size CT images are used for the machine learning. The Adam algorithm [13] with learning rate: 1*e−* 4 is used for stochastic optimization. The learning rate is dynamically reduced by using the ReduceLROnPlateau in the Keras Api during the learning. The results accuracy: 0.9902 and error: 0.0325 are obtained. The architecture of obtaining the mask images from the CT images by using BConvLSTM U-Net model is given in Fig. 1. The samples and obtained mask images by using BConvLSTM U-Net model are given in Fig. 2. The sample images after applying graph-cut image processing [10] are given in Fig. 3.

**Fig 1.**
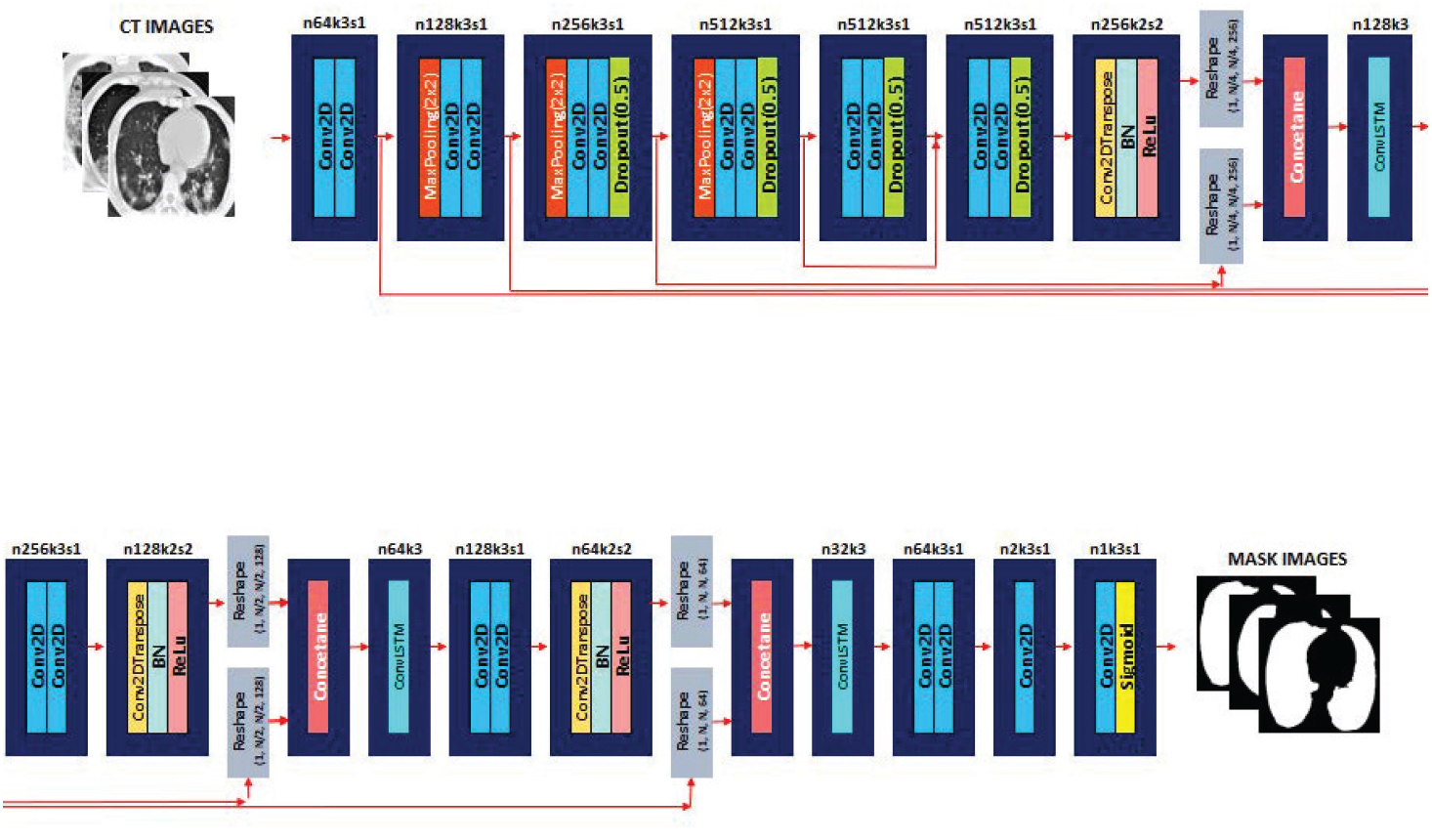
The architecture of obtaining the mask images from the CT images by using BConvLSTM U-Net model

**Fig 2.**
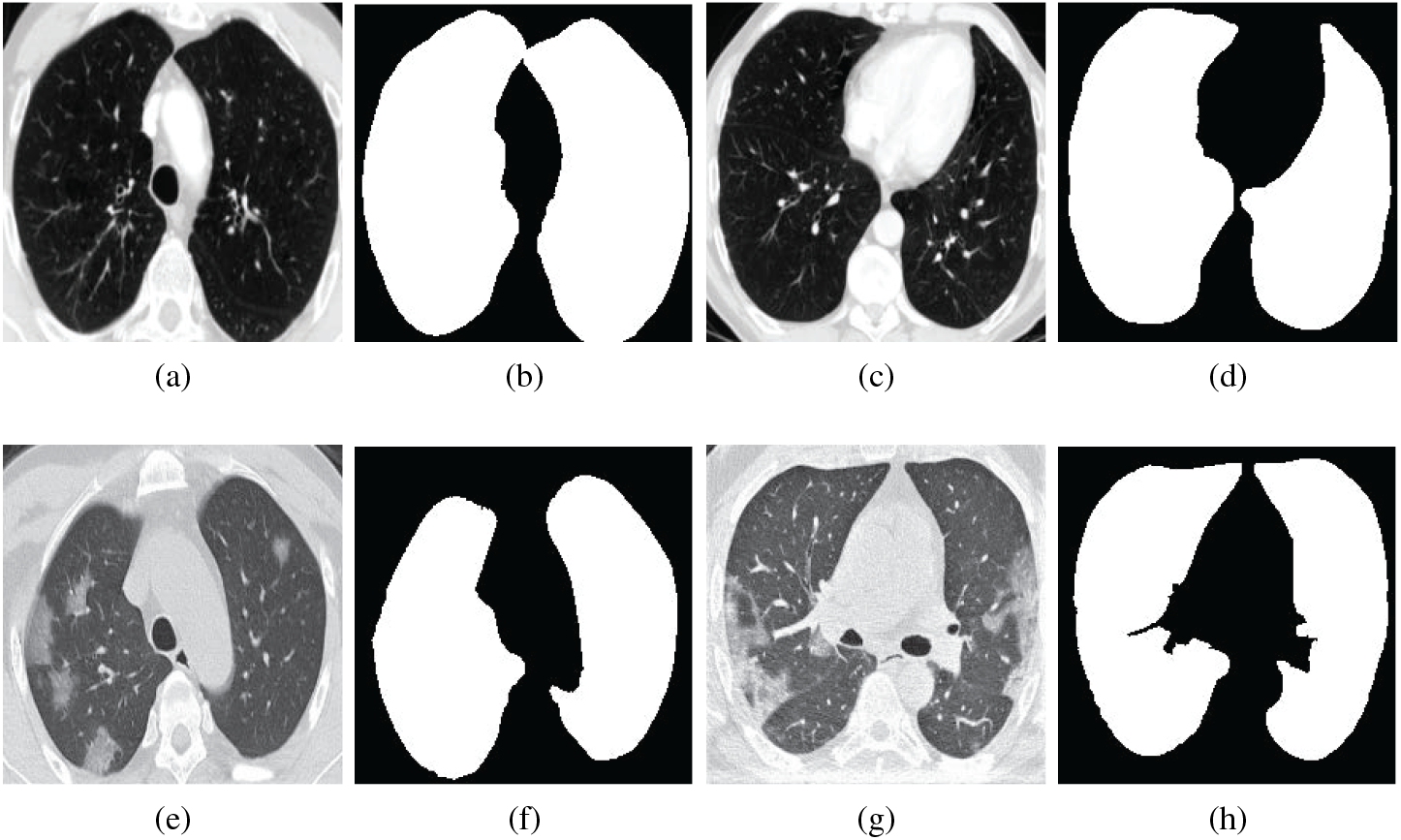
The samples and obtained mask images by using BConvLSTM U-Net model **a, c, e, g** Sample images **b, d, f, h** Mask images

**Fig 3.**
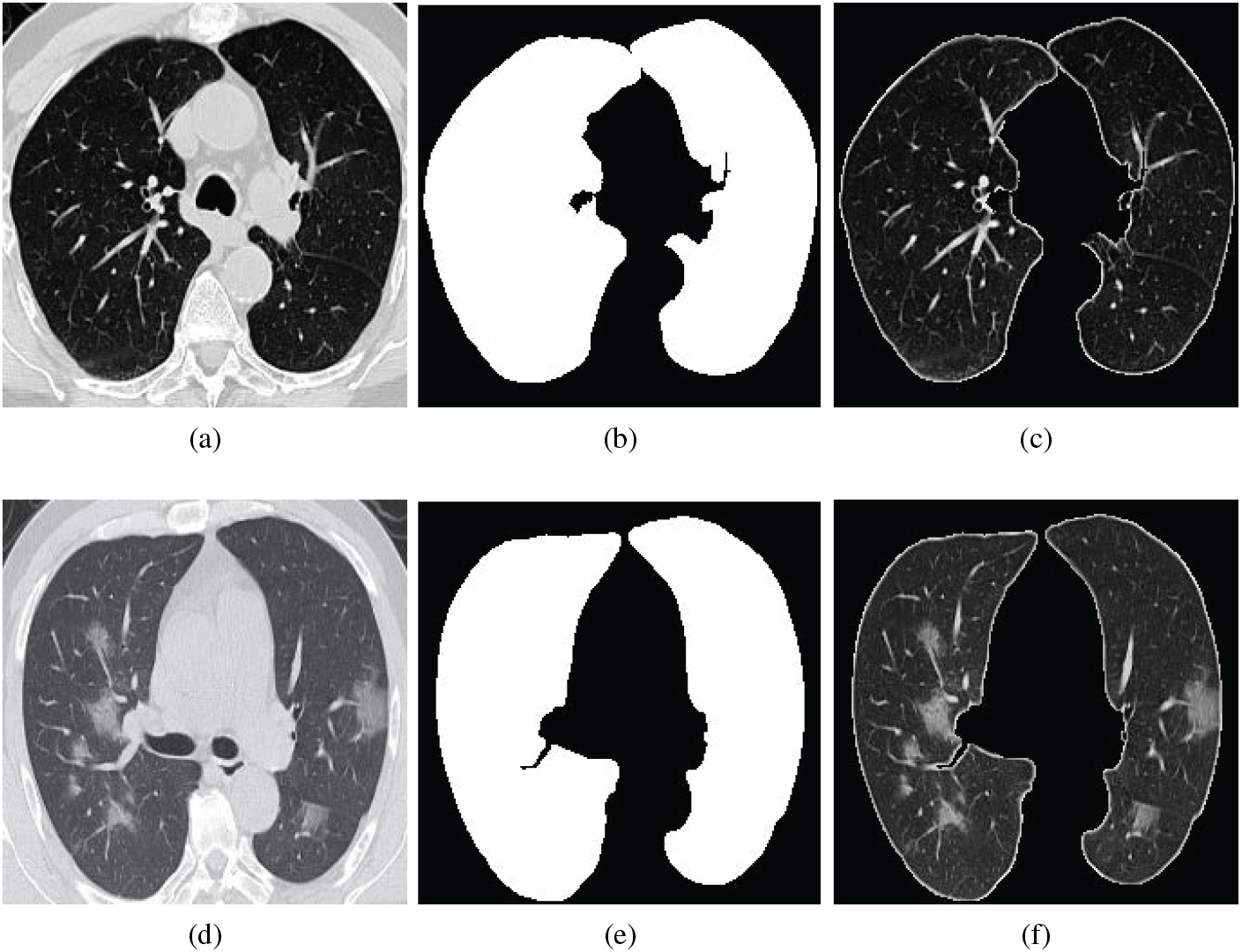
The sample images after applying graph-cut image processing **a, d** Sample images **b, e** Mask images **c, f** Graph-cut images

#### 2.2.2 Pre-processing

We use some pre-processing steps to optimize the training process in the training of deep learning models. These steps are resizing, image normalization and data augmentation. The images in the dataset vary in terms of resolution and size. The images of the relevant region are resized to 224 *×* 224 pixel. The intensity values of all images are normalized from [0, 255] to the standard normal distribution by min-max normalization to the intensity range of [0, 1] as follows.

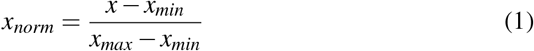

where *x* is the pixel intensity. *x*_*min*_ and *x*_*max*_ are minimum and maximum intensity values of the input image. This operation helps to speed up the convergence of the model by achieve a uniform distribution across the dataset. We use image data augmentation (DA) [14] for deep learning to improve the effectiveness of learning models. Random distortion, Flip (left, right), Rotate and Zoom types of data augmentation are applied to the images.

#### 2.2.3 Generative adversarial network (GAN)

Adam optimization algorithm with learning rate: 0.0001 is used for discriminator and generator training. The Least Squares function is used as the loss function in the study. The training epoch is 40000. 1232 real images with COVID-19 are used in the study. 3768 synthetic images with COVID-19 are produced as a result of the GAN training on real images with COVID-19 by using the generator. 1668 real healthy images are used in the study. 3332 synthetic healthy images are produced as a result of the GAN training on real healthy images by using the generator. The architecture and framework of GAN applied in the study are given in Figs. 4 and 5, respectively.The samples of the COVID-19 and healthy synthetic images produced after the GAN training are given in Figs. 6 and 7, respectively.

**Fig 4.**
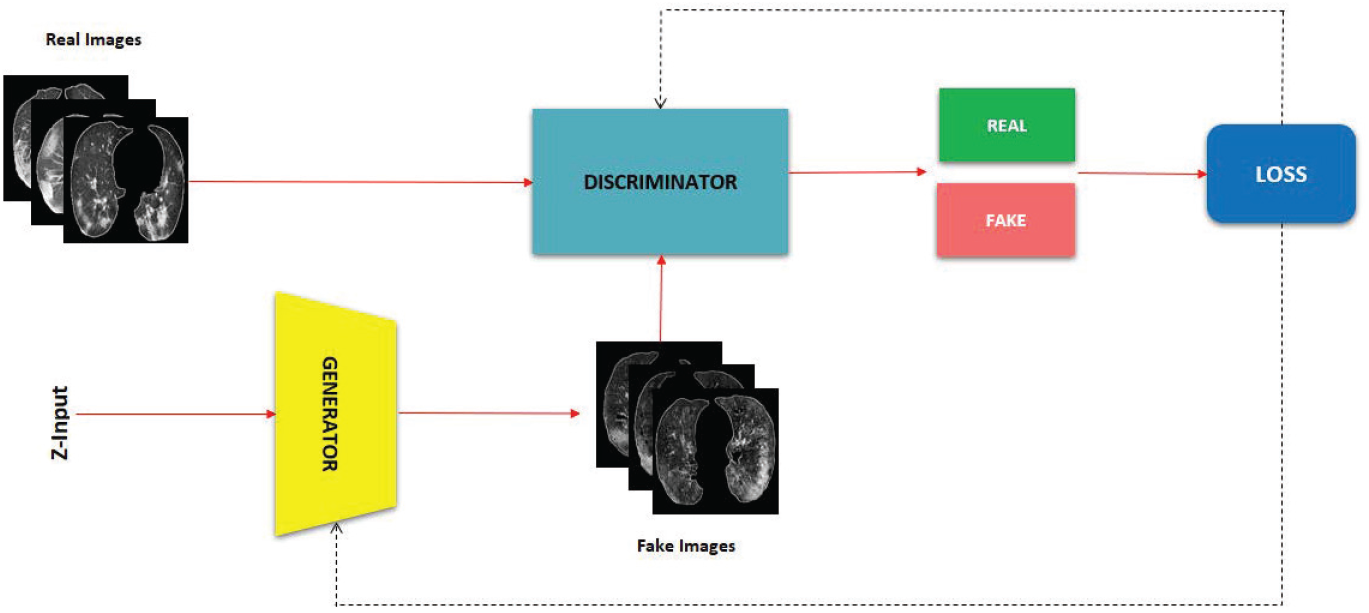
The architecture of GAN

**Fig 5.**
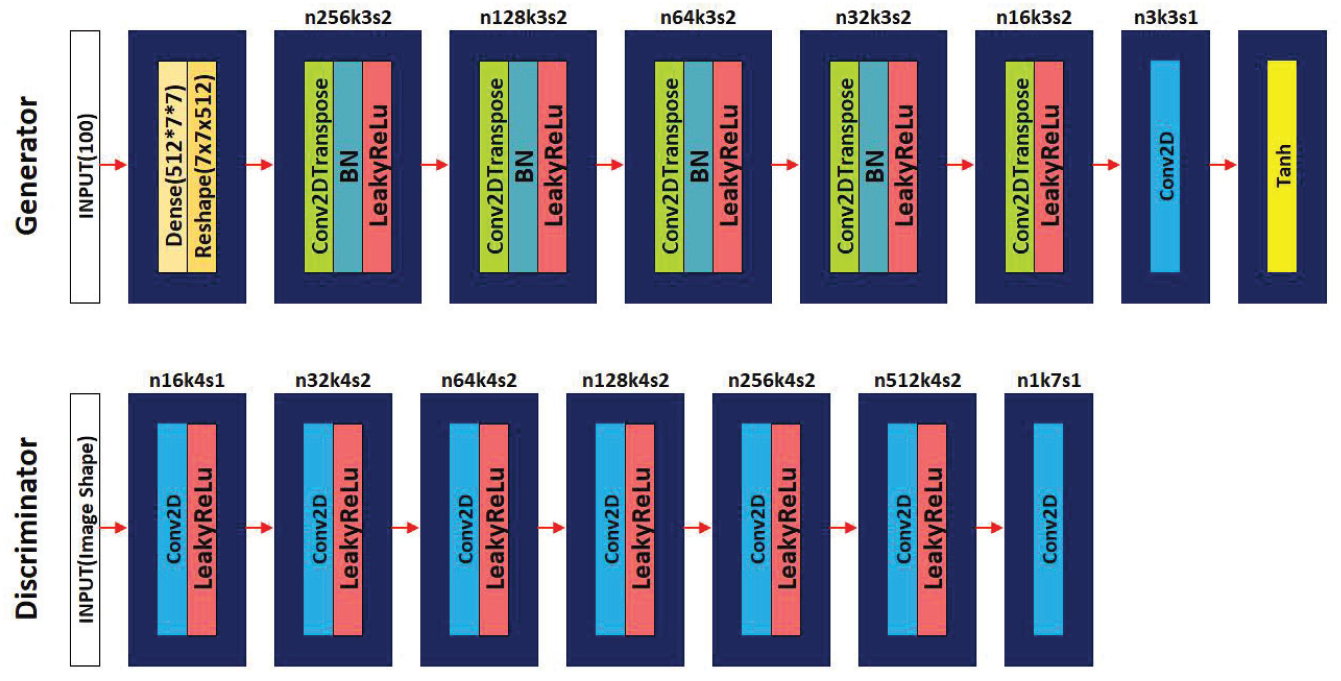
The framework of GAN

**Fig 6.**
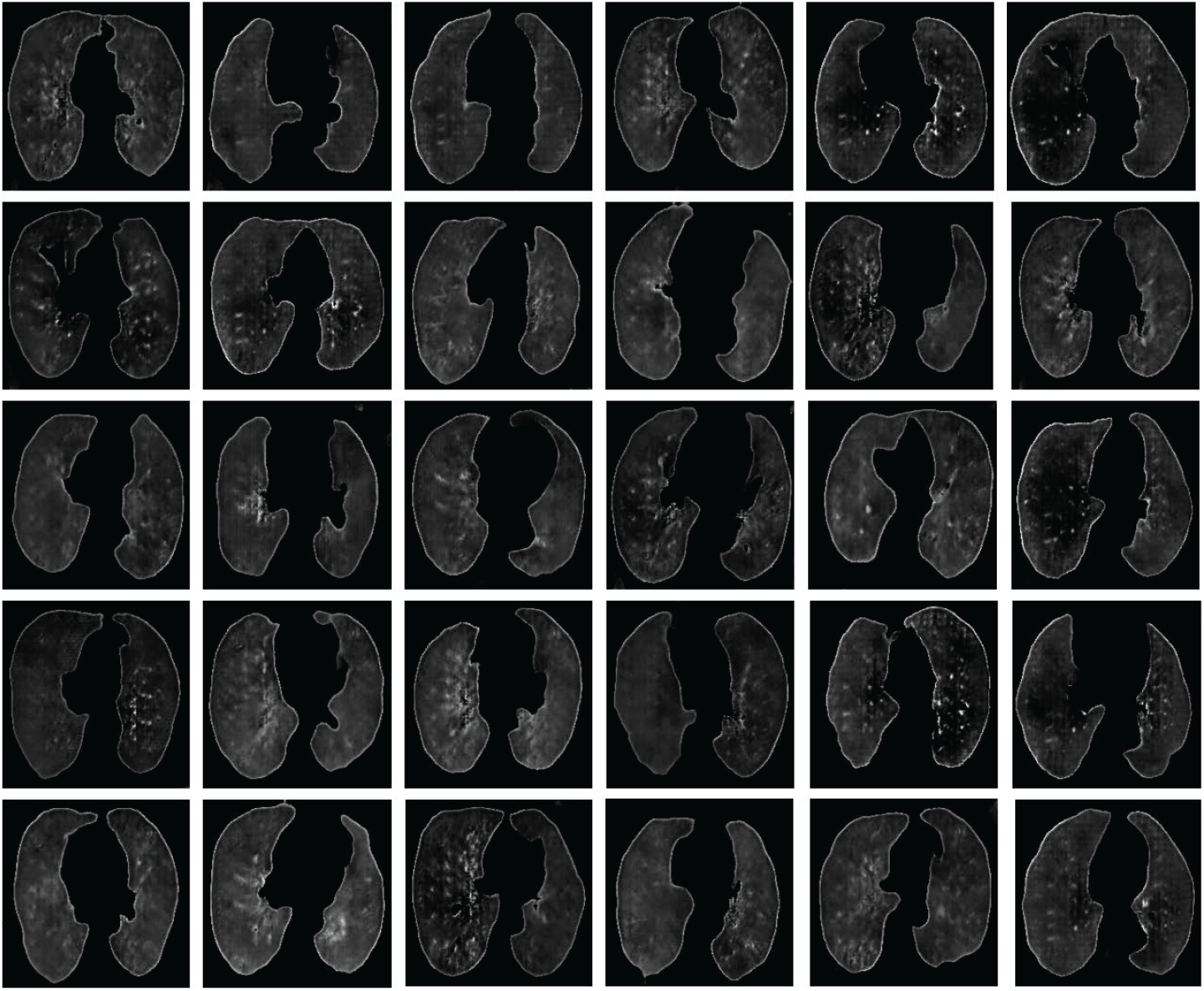
The samples of the COVID-19 synthetic images produced after the training without GAN

**Fig 7.**
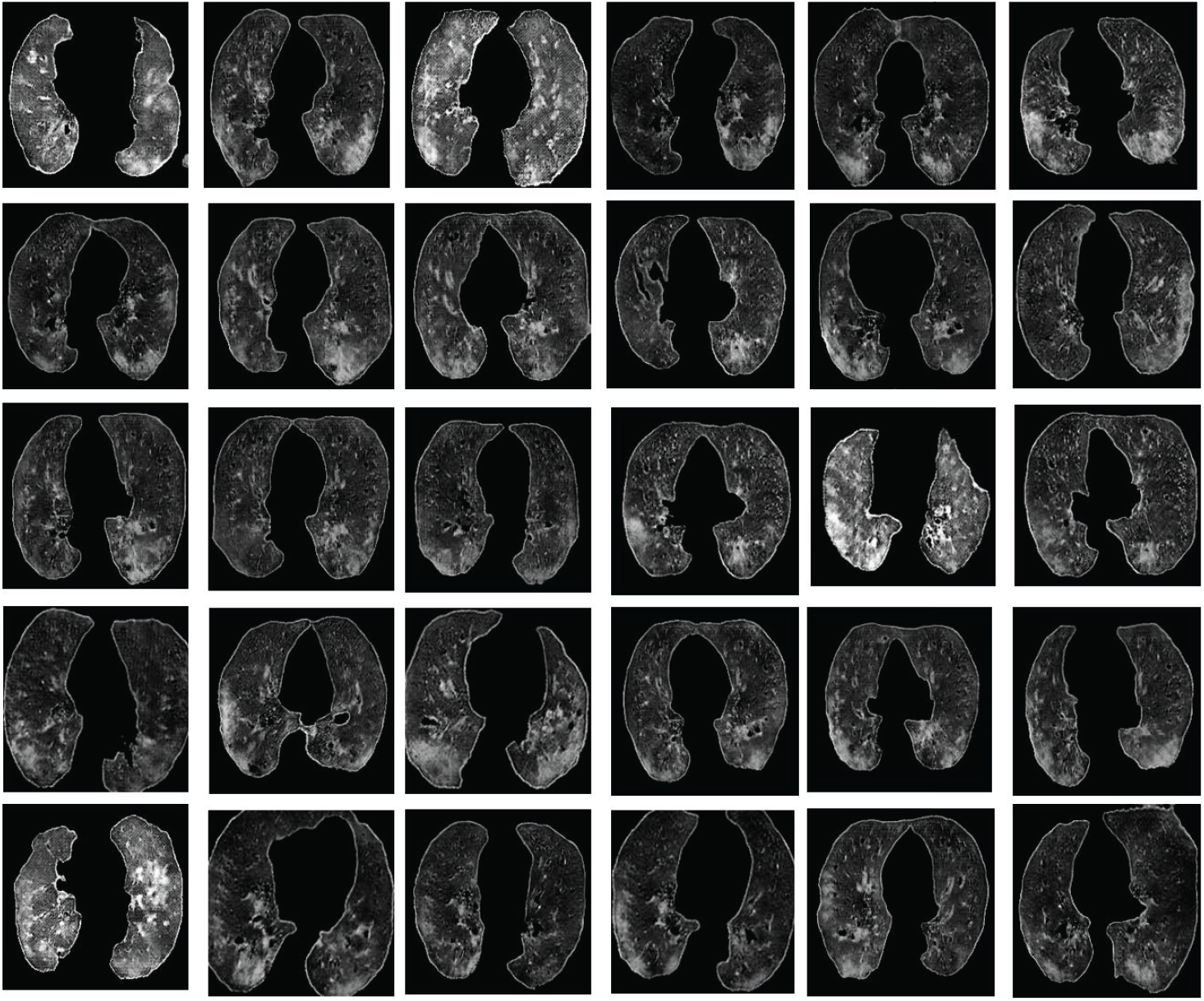
The samples of the healthy synthetic images produced after the training with GAN

### 2.3 Experimental evaluation

In this paper, two types of experimental studies are carried out in frameworks with and without (only with original samples) GAN. The frameworks without and with

GAN are given in Figs. 8 and 9, respectively. The numbers of the real images without GAN and the real and synthetic images produced with GAN in sets are shown in Table 1. In the framework with GAN, totally 7200 samples are used in the training set (including 3600 COVID-19 and 3600 healthy) are used to train the model by the validation set of totally 800 samples (400 COVID-19 and 400 healthy) and then test the model performance by the testing set of totally 2000 samples (1000 COVID-19 and 1000 healthy). To verify the validity of the operations, the data augmentation operations which are applied to both the original sets and the extended with GAN sets are shown in Table 2.

**Table 1.** The numbers of the real images without GAN and the real+synthetic images produced with GAN in training, validation and testing sets

| Framework | Types | Training set | Validation set | Testing set |
| --- | --- | --- | --- | --- |
| Real images without GAN | COVID-19 | 986 | 99 | 246 |
|  | Healthy | 1334 | 133 | 334 |
| Real+Synthetic images produced with GAN | COVID-19 | 3600 | 400 | 1000 |
|  | Healthy | 3600 | 400 | 1000 |

**Table 2.** Types of Data Augmentation

| Types | Parameters |
| --- | --- |
| Random distortion | probability=0.5<br>grid width=4<br>grid height=4 |
| Flip (left, right) | probability=0.5 |
| Rotate | probability=0.5<br>max left rotation=10<br>max right rotation=10 |
| Zoom | probability=0.5<br>min factor=0.9<br>max factor=1.05 |

**Fig 8.**
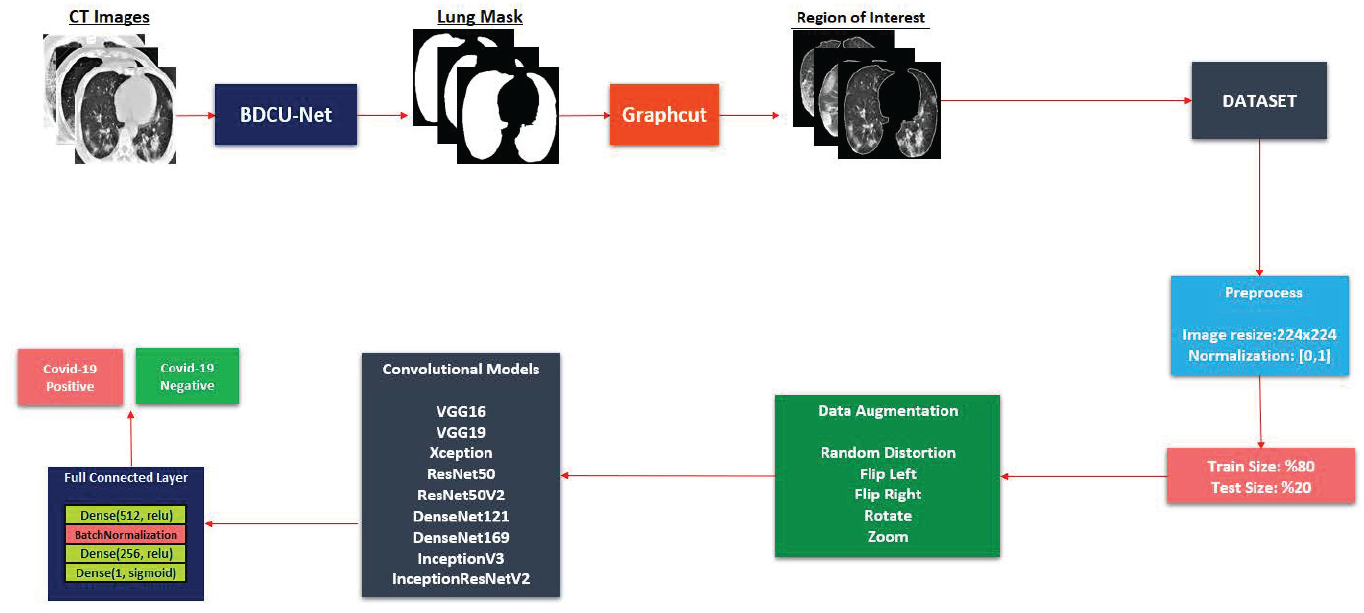
The framework without GAN

**Fig 9.**
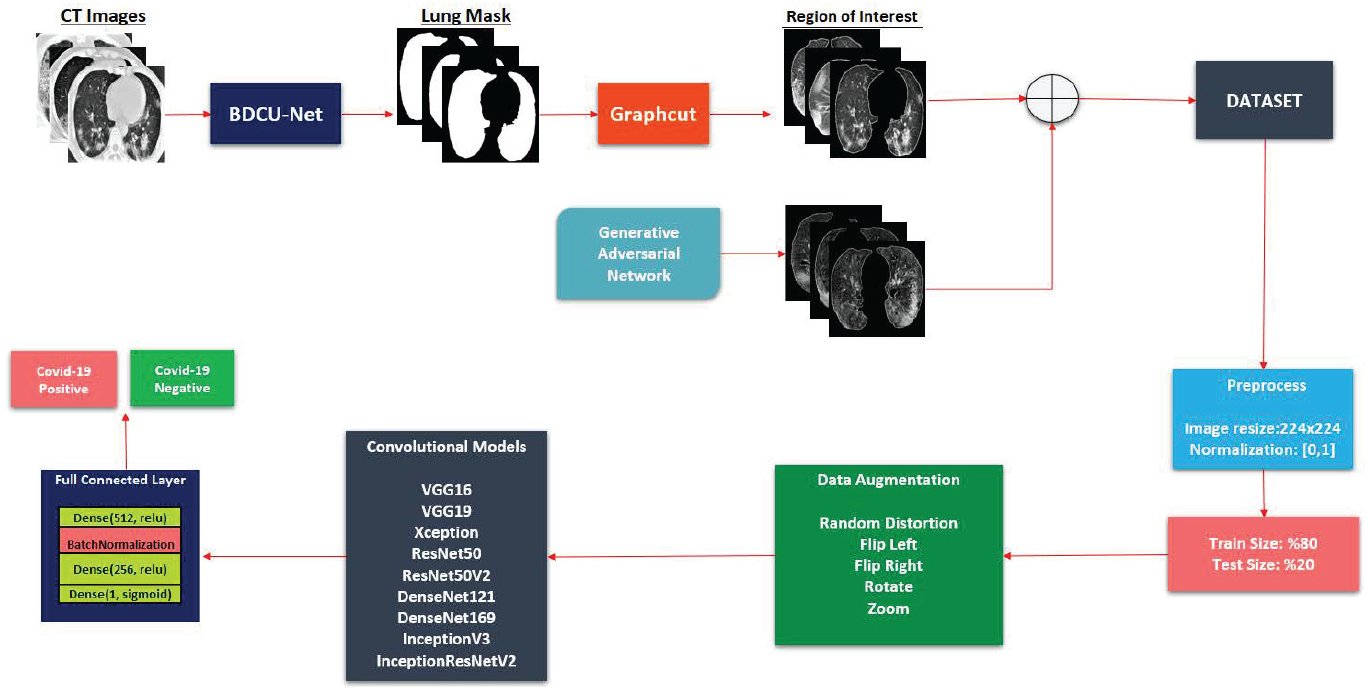
The framework with GAN

The methods followed in both frameworks are the same except for the datasets used. Keras Api is used for training, the Adam function is used for optimization in the both frameworks. The learning rate is started as 0.001. During the training, the learning rate is dynamically reduced by using the ReduceLROnPlateau in the Keras Api. The deep learning models, VGG16, VGG19, Xception, ResNet50, ResNet50V2, DenseNet121, DenseNet169, InceptionV3 and InceptionResNetV2 are used in the both frameworks. Dense(512, relu), BatchNormalization, Dense(256, relu), Dense (1, Sigmoid) layers are used in the full connected layer for classification, respectively.

## 3 Results

Estimation performances of the methods in this study are measured with metrics such as accuracy, precision, recall and f1-score. All evaluation metrics were calculated as follows according to Kassani et al. [2].

Dividing the number of correctly classified cases into the total number of test images shows the accuracy and is calculated as follows.

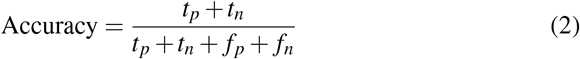

where *t*_*p*_ is the number of instances that correctly predicted, *f*_*p*_ is the number of instances that incorrectly predicted, *t*_*n*_ is the number of negative instances that correctly predicted, *f*_*n*_ is the number of negative instances that incorrectly predicted.

The recall is used to measure correctly classified COVID-19 cases. Recall is calculated as follows.

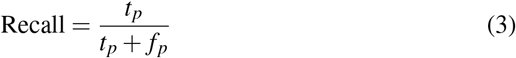

The percentage of correctly classified labels in truly positive patients is defined as the precision and is calculated as follows.

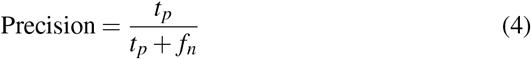

The F1-score is defined as the weighted average of precision and recall combining both precision and recall, and is calculated as follows.

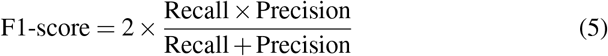

In the studies, the operations are performed on 334 healthy and 246 with COVID-19 images for the framework without GAN, and 1000 healthy and 1000 with COVID-19 images for the framework with GAN. The confusion matrices of the our framework without GAN and with GAN are given in Figs. 10 and 11, respectively. The comparative classification performance results of the deep learning models used in the our framework without GAN are shown in Table 3. Likewise, the results in the our framework with GAN are shown in Table 4.

**Table 3.** The comparative classification performance result of the deep learning models used in our framework without GAN

| Model | Accuracy | Error | Precision | Recall | F1-score | Roc Auc |
| --- | --- | --- | --- | --- | --- | --- |
| VGG16 | 0.974137 | 0.076263 | 0.967742 | 0.988024 | 0.977778 | 0.992941 |
| VGG19 | 0.968955 | 0.075962 | 0.981407 | 0.964072 | 0.972810 | 0.997574 |
| Xception | 0.991379 | 0.022388 | 0.991045 | 0.994012 | 0.992526 | 0.999769 |
| ResNet50 | 0.965517 | 0.1229 | 0.951149 | 0.991018 | 0.970674 | 0.985626 |
| ResNet50V2 | 0.981034 | 0.045709 | 0.973607 | 0.994012 | 0.983704 | 0.999148 |
| DenseNet121 | 0.982759 | 0.094841 | 0.982143 | 0.988024 | 0.985075 | 0.995211 |
| DenseNet169 | 0.982759 | 0.098201 | 0.976471 | 0.994012 | 0.985163 | 0.989661 |
| InceptionV3 | 0.993103 | 0.022390 | 0.994012 | 0.994012 | 0.994012 | 0.999647 |
| InceptionResNetV2 | 0.989655 | 0.064712 | 0.985207 | 0.997006 | 0.991071 | 0.998406 |

**Table 4.**
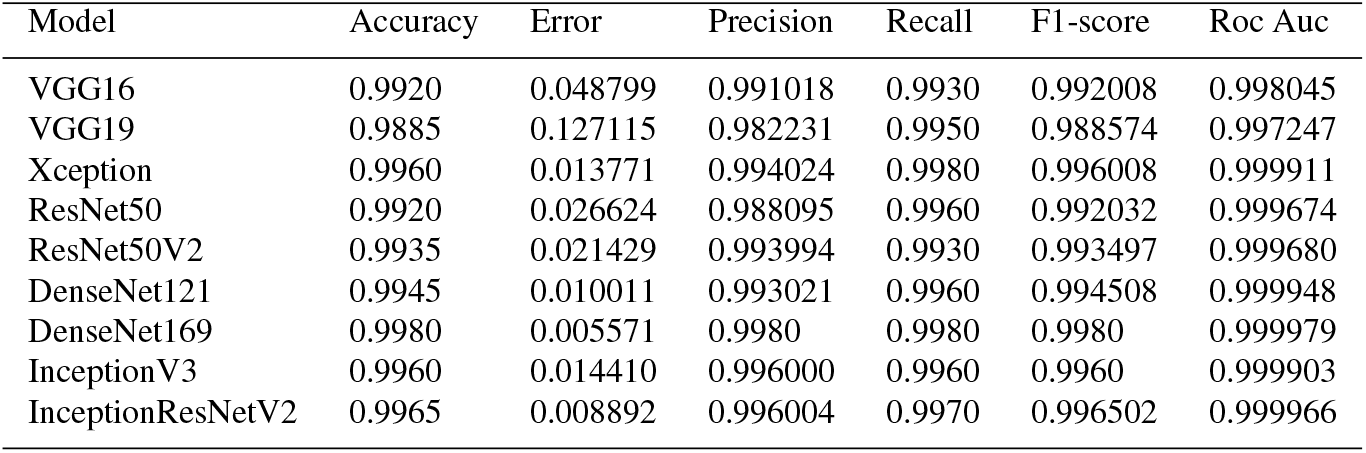
The comparative classification performance result of the deep learning models used in our frame-work with GAN

**Fig 10.**
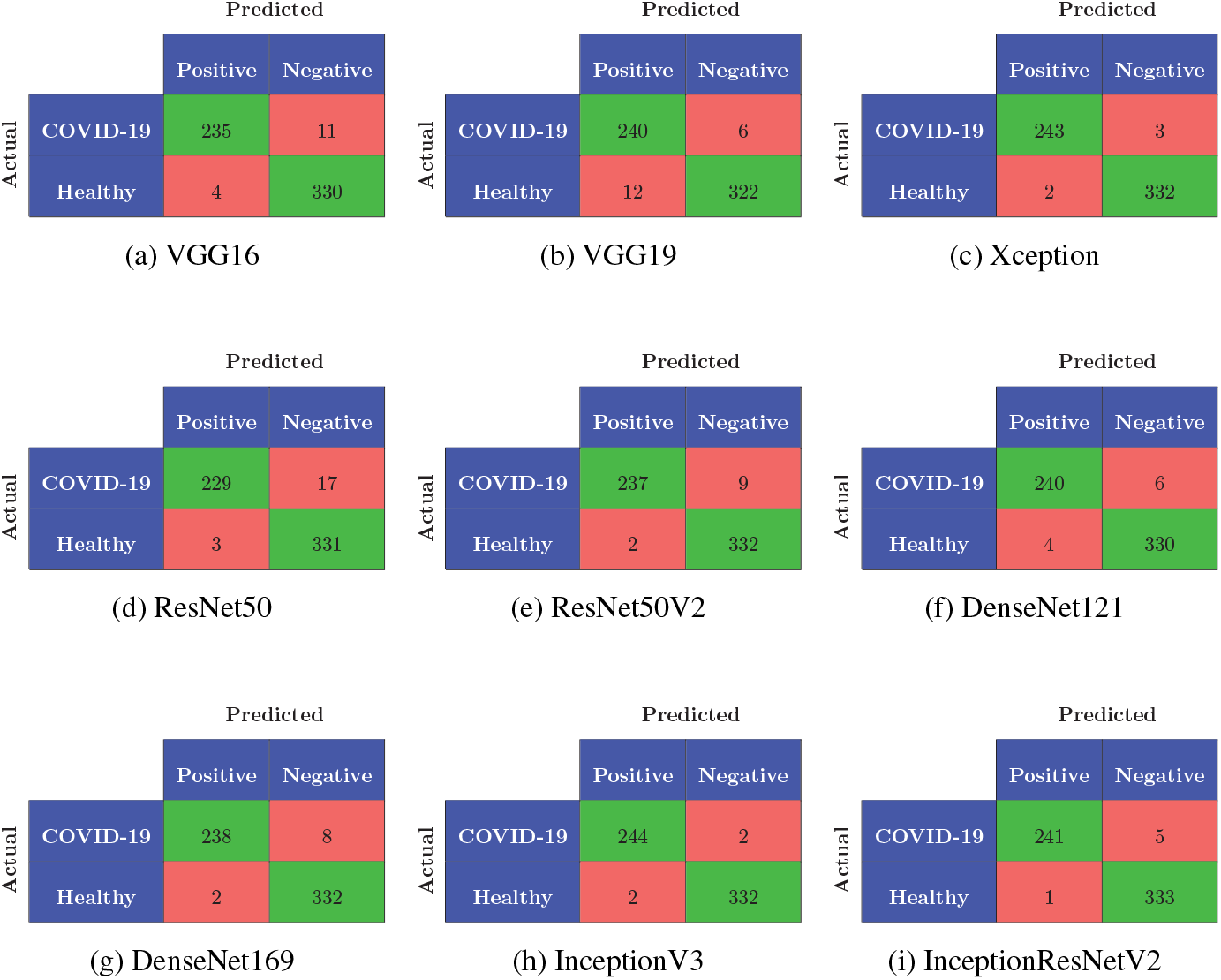
The confusion matrices of the our framework without GAN

**Fig 11.**
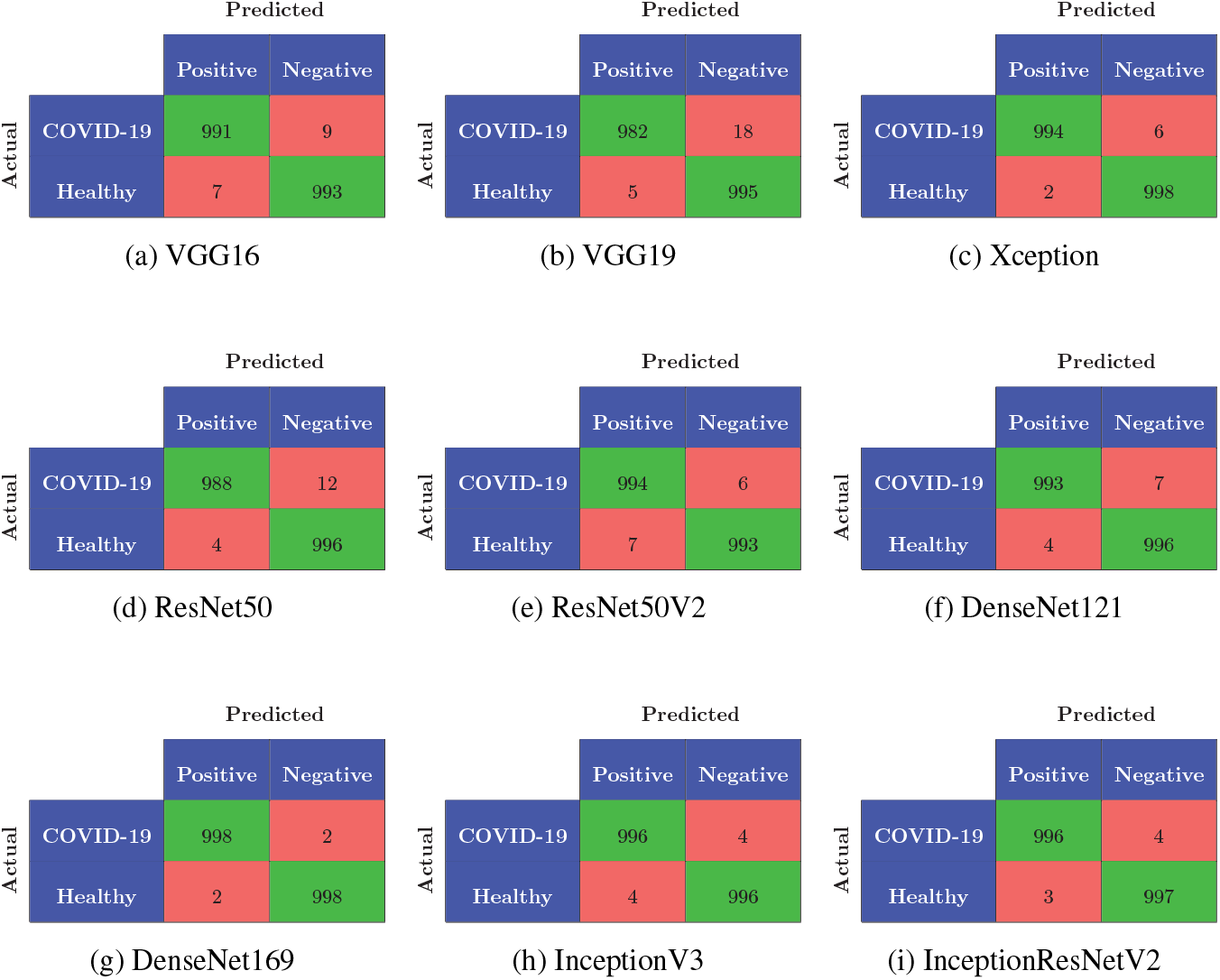
The confusion matrices of the our framework with GAN

## 4 Discussion

The imaging methods, such as CT, play a more important role in making the initial diagnosis for COVID-19 than the Polymerase Chain Reaction (PCR) tests. The performance of deep learning models depends directly on the quality of the features determined from the image. The focus of this study, which is based on the success of deep learning models, is to achieve high accuracy results using features related to the classification of COVID-19 in CT imaging. The machine is trained by using BConvLSTM U-Net architecture for lung segmentation. In every experiment, the technique of reproducing original images with the GAN method is used to evaluate the performance of the classifiers.

Performance information of deep learning models such as VGG16, VGG19, Xception, ResNet50, ResNet50V2, DenseNet121, DenseNet169, InceptionV3 and InceptionResNetV2 without GAN and with GAN are shown in Tables 1and 2, respectively. In our studies, the highest accuracy rate is 99.80% with the DenseNet169 method, the highest precision rate is 99.80% with the DenseNet169 method, the highest recall rate is 99.80% with the DenseNet169 method, the highest f1-score is 99.80% with the DenseNet169 method and the highest roc acu is 99.9979% with the DenseNet169 method. All of the highest values are obtained with the DenseNet169 method. Therefore, we can say that the DenseNet169 method is more efficient than the other deep learning methods.

The image segmentation methods and the best accuracy results of the studies in COVID-19 applications with CT images and our method are shown in Table 5. The high number of samples used in deep learning and the segmentation architecture used are very important for higher accuracy. In our study, more samples are used than the existing studies.

**Table 5.** The image segmentation methods and the best accuracy results of the studies in COVID-19 applications with CT images

| Literature | Images | Method | Accuracy(%) |
| --- | --- | --- | --- |
| Our proposed with GAN | 1000 COVID-19<br>1000 Others | BConvLSTM<br>U-Net | 99.80 |
| Chen et al. [3] | 51 COVID-19<br>55 Others | UNet++ | 95.2 |
| Zheng et al. [4] | 313 COVID-19<br>229 Others | U-Net | 90.8 |
| Wang et al. [15] | 44 COVID-19<br>55 Others | CNN | 82.9 |

It is clearly seen from the Table 5 that we obtained the highest accuracy value 99.80% in the literature along with the BConvLSTM U-Net method that we use for image segmentation as well as many examples.

In addition, Kassani et al. [2] perform accuracy, precision, recall and f1-score analyzes with different deep learning algorithms without image segmentation. The accuracy, precision, recall and f1-score analyzes with different deep learning algorithms in the our and Kassani et al.’s [2] frameworks are shown in Table 6.

**Table 6.** The accuracy, precision, recall and f1-score analyzes with different deep learning algorithms in the our and Kassani et al.’s [2] frameworks

| Model | Framework | Accuracy(%) | Precision(%) | Recall(%) | F1-score(%) |
| --- | --- | --- | --- | --- | --- |
| VGG16 | Our proposed | 99.20 | 99.1018 | 99.30 | 99.2008 |
|  | Ref. [2] | 91.00 | 94.00 | 94.00 | 94.00 |
| VGG19 | Our proposed | 98.85 | 98.2231 | 99.50 | 98.8574 |
|  | Ref. [2] | 90.00 | 94.00 | 94.00 | 94.00 |
| Xception | Our proposed | 99.60 | 99.4024 | 99.80 | 99.6008 |
|  | Ref. [2] | 96.00 | 98.00 | 98.00 | 98.00 |
| ResNet50 | Our proposed | 99.20 | 98.8095 | 99.60 | 99.2032 |
|  | Ref. [2] | 98.00 | 96.00 | 96.00 | 96.00 |
| ResNet50V2 | Our proposed | 99.35 | 99.3994 | 99.30 | 99.3497 |
|  | Ref. [2] | 96.00 | 96.00 | 96.00 | 96.00 |
| DenseNet121 | Our proposed | 99.45 | 99.3021 | 99.60 | 99.4508 |
|  | Ref. [2] | 99.00 | 98.00 | 98.00 | 98.00 |
| InceptionV3 | Our proposed | 99.60 | 99.60 | 99.60 | 99.60 |
|  | Ref. [2] | 95.00 | 99.00 | 99.00 | 99.00 |
| InceptionResNetV2 | Our proposed | 99.65 | 99.6004 | 99.70 | 99.6502 |
|  | Ref. [2] | 94.00 | 96.00 | 95.00 | 95.00 |

It can be clearly seen from Table 6 that our framework is more successful than the Kassani et al.’s framework [2] in terms of accuracy, precision, recall and f1-score rates in all of the deep learning algorithms.

In conclusion, the highest available rates are achieved in terms of accuracy (99.80%), precision (99.80%), recall (99.80%), f1-score (99.80%) and roc acu (99.9979%) with deep learning methods in this paper. The our presented framework with GAN in this paper is more successful than all the existing studies that classify COVID-19 with the deep learning algorithms.

## Data Availability

We regard the availability of all data referred to in the manuscript.

## Availability of data and material

Not applicable

## Code availability

Not applicable

## Authors’ contributions

All authors contributed to the study conception and design.

Conceptualization: Erdi Acar, Engin Ş ahin, İhsan Yilmaz; Data curation: Erdi Acar; Formal analysis: Erdi Acar, Engin Ş ahin; Investigation: Erdi Acar, Engin Ş ahin, İhsan Yilmaz; Methodology: Erdi Acar, İhsan Yilmaz; Resources: Erdi Acar; Software: Erdi Acar; Supervision: İhsan Yilmaz; Visualization: Engin Ş ahin; Writing - original draft preparation: Engin Ş ahin; Writing - review and editing: Engin Ş ahin;

All authors read and approved the final manuscript.

